# A GWAS for grip strength in cohorts of children – advantages of analysing young participants for this trait

**DOI:** 10.1101/2024.02.09.24302539

**Authors:** Filippo Abbondanza, Carol A. Wang, Judith Schmitz, Craig E. Pennell, Andrew J.O. Whitehouse, Silvia Paracchini

## Abstract

Grip strength (GS) is a proxy measure for muscular strength and a predictor for bone fracture risk among other diseases. Previous genome-wide association studies (GWAS) have been conducted in large cohorts of adults focusing on scores collected for the dominant hand, therefore increasing the likelihood of confounding effects by environmental factors. Here, we perform the first GWAS meta-analyses on maximal GS with the dominant (GSD) and non-dominant (GSND) hand in two cohorts of children (ALSPAC, N = 5,450; age range = 10.65 – 13.61; Raine Study, N = 1,162, age range: 9.42-12.38 years).

We identified a novel significant association for GSND (rs9546244, *LINC02465*, *p* = 3.43e-08) and replicated associations previously reported in adults including with a *HOXB3* gene marker that shows an eQTL effect. Despite a much smaller sample (∼3%) compared to the UK Biobank we replicated correlation and polygenic risk score (PRS) analyses previously reported in this much larger adult cohort. Specifically, we observed genetic correlations with coronary artery disease and a PRS association with the risk of overall fracture. Furthermore, we observed a higher SNP-heritability (24-41%) compared to previous studies (4-24%) in adults. Our results suggest that cohorts of children might be better suited for genetic studies of grip strength, possibly due to the shorter exposure to confounding environmental factors compared to adults.

## Introduction

Grip strength (GS) is an indicator of muscle fitness and has been shown to predict different health outcomes (Ortega et al., 2012; Smith et al., 2019; Taekema et al., 2010). In a longitudinal study on postmenopausal women (N = 2,928) low GS – measured for the dominant hand – was associated with a slight increase of hip fractures (OR = 1.05, 95% CI 1.01-1.09) (Kärkkäinen et al., 2008). Cheung et al. reported similar findings, where low GS scores (< 1 SD from the mean) were associated with increased risk of major fractures in the Hong Kong population (OR = 1.24, 95% CI 1.09-1.42; N = 4,855) (Cheung et al., 2012). A longitudinal study on more than 1 million males found that GS was a reliable predictor of premature mortality, with stronger individuals presenting a lower risk for cardiovascular diseases, suicide, and psychiatric disorders (Ortega et al., 2012). Analysis in the UK Biobank found supportive evidence of a protective role of relative GS (calculated as the average mean of the right– and left-hand measurements divided by body weight) for atrial fibrillation (OR = 0.75, 95% CI 0.62–0.90) and coronary artery disease (CAD, OR = 0.69, 95% CI 0.60–0.79) (Tikkanen et al., 2018). The authors also reported a strong negative genetic correlation between GS, depressive symptoms and attention deficit hyperactivity disorder (ADHD). A separate study in the UK Biobank showed that maximal grip strength is positively associated with cognitive measures of reasoning, prospective memory, number memory, visual memory, and reaction time (Firth et al., 2018).

In twin studies, GS heritability has been estimated in the range of 30-65% (Arden & Spector, 1997; Matteini et al., 2016). To date, few studies have investigated the molecular genetics of GS. An initial genome-wide association study (GWAS) conducted in two small Australian cohorts (N_total_ = 2,629, age range 55-85) found no variants associated with maximal GS for the dominant hand (Chan et al., 2015). Analysis in the UK Biobank (N = 195,180, age range 40-69) identified 16 statistically significant associations and reported the SNP-heritability (SNP-h^2^) for maximal grip strength to be 24% (Willems et al., 2017). Statistically significant associations were located close to genes known to have a neuromuscular function (*ACTG1*) or implicated in rare monogenic syndromes characterised by progressive neurological/psychomotor impairment (*KANSL1* and *LRPPRC)*. Associations were also detected in *HOXB3*, a highly conserved gene essential for morphogenesis and the establishment of the anterior-posterior axis.

A second GWAS in the UK Biobank (N = 223,315) on relative GS identified 64 associated markers (Tikkanen et al., 2018). The lead variants were located close to genes involved in muscular development disease (myotonic dystrophy type 1, Pitt-Hopkins syndrome, congenital heart defect and hand abnormalities, Char syndrome) and intellectual disability with a variant located in a long non-coding RNA (*LINC01874*).

A recent meta-analysis of categorical GWAS for GS (low vs high) across 22 cohorts of elderly individuals (total N = 256,523 individuals aged 60 years and over, including 200,565 UK Biobank participants), identified 15 associated loci (Jones et al., 2021). Associated genes are involved in autoimmune disease (*HLA-DQA1*) and osteoarthritis (*GDF5, ALDH1A2, SLC39A8*). Significant genetic correlations were found between low GS and osteoarthritis (r_g_ = 0.30), CAD (r_g_ = 0.19) and Alzheimer (r_g_ = 0.16). Exome sequencing analysis in the UK Biobank identified rare variants associated with GS in the *KDM5B*, *OBSCN*, *GIGYF1*, *TTN*, *RB1CC1*, and *EIF3J* genes (Huang et al., 2023).

Diet, sports activities, and health conditions are expected to contribute to GS, potentially masking genetic effects. For example, analyses in younger participants, who accumulated less environmental exposures, might be more effective in identifying genes contributing to GS. Furthermore, we reasoned that the non-dominant hand would be less influenced by training from everyday activities and therefore more likely to reveal genetic effects. Therefore, we conducted the analysis for grip strength testing both the dominant (GSD) and non-dominant (GSND) hands. We report the first GWAS for two GS measures conducted in two cohorts of children (ALSPAC: N = 5,450, mean age = 11.8 years; Raine Study: N = 1,162, mean age = 10.6 years). Genetic correlation and polygenic risk score (PRS) analyses were performed for phenotypes, such as cardiovascular disorders, heel bone density and psychiatric disorders previously reported to be associated with GS in both behavioural and genetic levels.

## Methods

### Cohorts

#### ALSPAC cohort

Pregnant women resident in Avon, UK with expected dates of delivery between 1st April 1991 and 31st December 1992 were invited to take part in the Avon Longitudinal Study of Parents and Children (ALSPAC) (Boyd et al., 2013; Fraser et al., 2013). The initial number of pregnancies enrolled is 14,541. When the oldest children were approximately 7 years of age, an attempt was made to bolster the initial sample with eligible cases who had failed to join the study originally. The total sample size for analyses using any data collected after the age of seven is therefore 15,447 pregnancies. From age 7, all children were invited annually for assessments on a wide range of physical, behavioural and neuropsychological traits. Informed consent for the use of data collected via questionnaires and clinics was obtained from participants following the recommendations of the ALSPAC Ethics and Law Committee at the time. Consent for biological samples has been collected in accordance with the Human Tissue Act (2004). Ethical approval for the present study was obtained from the ALSPAC Law and Ethics Committee and the Local Research Ethics Committees.

Grip strength was assessed at age 11 using the Jamar hand dynamometer, which measures isometric strength in kilograms. Before starting the test, the children were instructed on how to use the instrument and were given a practice trial. Children were encouraged to squeeze the dynamometer as long as possible. The test was repeated for each hand separately three times. The absolute highest score across all trials determined grip strength with the dominant hand (GSD). The maximal score for the other hand determined grip strength with the non-dominant hand (GSND). For the current study, we removed individuals with missing phenotype or genotype data and outliers ( outliers (N < 5 with very high grip strength scores of 65kg and 42kg), resulting in a total of 5,450 participants entering the analyses (mean age = 11.8 years, SD = 0.24, N_males_ = 2,674, N_Females_ = 2,776; Table 1).

**Table 1.** Grip strength mean.

| Sex | ALSPAC (SD) in kg |  |  | Raine Study (SD) in kg |  |  |
| --- | --- | --- | --- | --- | --- | --- |
|  | N | GSD | GSND | N | GSD | GSND |
| Male | 2,674 | 20.8 (4.32) | 18.7 (4.08) | 609 | 17.0 (3.21) | 15.17 (3.14) |
| Female | 2,776 | 19.6 (4.19) | 17.6 (4.03) | 553 | 15.44 (3.12) | 13.59 (3.01) |
| Combined | 5,450 | 20.2 (4.29) | 18.2 (4.09) | 1,162 | 16.26 (3.26) | 14.42 (3.18) |

#### Raine Study

The Raine Study is a prospective pregnancy cohort of 2900 mothers recruited between 1989–1991 (https://rainestudy.org.au/) (Newnham et al., 1993). Recruitment took place at Western Australia’s major perinatal centre, King Edward Memorial Hospital, and nearby private practices. Women who had sufficient English language skills, an expectation to deliver at King Edward Memorial Hospital, and an intention to reside in Western Australia to allow for future follow-up of their child were eligible for the study.

The primary carers (Gen1) completed questionnaires regarding their respective study child, and the children (Gen2) had physical examinations at ages 1, 2, 3, 5, 8, 10, 14, 17, 18, 20, and 22. Ethics approval for the original pregnancy cohort and subsequent follow-ups were granted by the Human Research Ethics Committee of King Edward Memorial Hospital, Princess Margaret Hospital, the University of Western Australia, and the Health Department of Western Australia. Parents, guardians and young adult participants provided written informed consent either before enrolment or at data collection at each follow-up. All research was performed in accordance with the approved guidelines.

Grip strength was assessed at Raine Study Gen2 10 follow-up using the as part of the psychometric properties of the US developed McCarron Assessment of Neuromuscular Development (MAND) (McCarron, 1997). Grip strength was measured with a hand dynamometer, alternated twice on each hand. The skilled research assistant administering the assessment first demonstrated the task and instruction to “squeeze the handle as hard as you can” was given prior to each attempt. Raw scores (measured in kilograms) were converted to scaled scores based on the participant’s age. The absolute highest score across all trials determined grip strength with the dominant hand (GSD). The maximal score for the other hand determined grip strength with the non-dominant hand (GSND). For the current study, we removed individuals with missing phenotype or genotype data, resulting in a total of 1,162 participants entering the analyses (mean age = 10.59 years, SD = 0.18, N_males_ = 609, N_Females_ = 553; Table 1).

## Genotyping, quality control and imputation

### ALSPAC

Standard quality control (QC) and imputations were performed centrally by the ALSPAC team. Briefly, participants were genotyped using the Illumina HumanHap550 quad chip. The resulting raw genome-wide data were subjected to standard QC methods. Individuals were excluded on the basis of gender mismatches; minimal or excessive heterozygosity and disproportionate levels of individual missingness (>3%). Population stratification was assessed by multidimensional scaling analysis and compared with HapMap II (release 22) European descent (CEU), Han Chinese, Japanese and Yoruba reference populations; all individuals with non-European ancestry were removed. SNPs with a minor allele frequency (MAF) of < 1%, a call rate of < 95% or evidence for violations of Hardy-Weinberg equilibrium (*p* < 5e-07) were removed. Cryptic relatedness was measured as the proportion of identity by descent (IBD > 0.1). Related subjects that passed all other quality control thresholds were retained during subsequent phasing and imputation. As a result, 9,115 subjects and 500,527 SNPs passed these quality control filters and were imputed using Impute v3 and the HRC 1.1 reference panel.

After imputation, SNPs with info score ≥ 0.4 and MAF ≥ 0.05 were retained for GWAS using BOLT-LMM, leaving a total of 5,405,480 SNPs.

### The Raine Study

The Raine Study Gen2 participants were genotyped on an Illumina 660 W Quad Array at the Centre for Applied Genomics, Toronto, Canada. QC of the GWAS genotyped data were performed as per standard protocol. In brief, a total of 1593 Raine Study Gen2 participants were genotyped on an Illumina 660 Quad Array, which included 657,366 genetic variants, consisting of ∼ 560,000 single-nucleotide-polymorphisms (SNPs) and ∼ 95,000 copy number variants (CNVs), at the Centre for Applied Genomics, Toronto, Canada. Plate controls and replicates with a higher proportion of missing data were excluded before individuals were assessed for low genotyping success (> 3% missing), excessive heterozygosity, gender discrepancies between the core data and genotyped data, and cryptic relatedness (π > 0.1875, in between second– and third-degree relatives—e.g. between half-siblings and cousins). At the SNP level, the SNP data were cleaned using plink (Purcell et al., 2007) following the Wellcome Trust Case—Control Consortium protocol (Wellcome Trust Case Control et al., 2007). The exclusion criteria for SNPs included: Hardy–Weinberg-Equilibrium *p* < 5.7 × 10–7; call-rate < 95%; MAF < 1%; and SNPs of possible strand ambiguity (i.e. A/T and C/G SNPs). The cleaned GWAS data were imputed using MACH software (Howie et al., 2012) across the 22-autosomes and X-chromosome against the 1000 Genome Project Phase I version 3 (Auton et al., 2015). A total of 1494 individuals with 535,632 SNPs remained after genotype QC; imputation resulted in 30,061,896 and 1,264,4493 SNPs across the 22-autosomes and X-chromosome, respectively. Principal components (PCs) analysis was carried out, using SMARTPCA from v.3.0 of EIGENSOFT (Price et al., 2006), on the cleaned genotyped data, where PCs were generated for purposes of adjusting for population stratification in all genetic analyses.

## Heritability estimates and GWAS

The primary analysis was conducted in ALSPAC because of the sufficiently large sample size. We computed univariate SNP-h^2^ with BOLT-REML, which leverages raw data, implemented in BOLT-LMM v2.3.4 (Loh, Bhatia, et al., 2015; Loh, Tucker, et al., 2015) using sex and age as covariates. Heritability in ALSPAC and the meta-analysed GWAS results were also estimated with LDSC (Bulik-Sullivan et al., 2015) for direct comparisons with previous studies.

GWAS analysis was conducted in ALSPAC (N = 5,450) with BOLT-LMM v2.3.4 (Loh, Tucker, et al., 2015) under the standard infinitesimal linear mixed model (LMM) framework. As BOLT-LMM implements linear mixed models, adjusting for PC was not needed. T

GWAS analysis was conducted in Raine Study (N = 1,162) with Probabel v0.4.1 (Aulchenko et al., 2010) under the standard linear model framework including the first two PCs as covariates. In both studies, sex and age were used as covariates on the basis of previously reported effects in behavioural analysis for grip strength conducted in the ALSPAC cohort (Buenaventura Castillo et al., 2020). The genomic inflation factor (λ) was calculated with LDSC.

The results from the separate GWAS were meta-analysed under inverse-variance fixed-analysis using METAL (Willer et al., 2010). Only markers available in both GWAS results were retained and were uploaded to FUMA to i) annotate the data using CADD v1.4, ii) identify markers previously reported in GWAS Catalogue, iii) create regional plots for significant hits, iv) run gene-set enrichment analysis using MAGMA v1.6 and MsigDB v6.2, v) identify possible eQTLs for top markers (*p* < 1e-04) using GTEx v8, and vi) run gene-based association using MAGMA v1.6 (de Leeuw et al., 2015). All results are reported in GRCh37 genome build.

### Replication analysis

The results from the meta-analyses were used to test for replication of 93 SNPs associated with GS in the previous literature (*p* < 5e-08), which were selected as predefined lead SNPs in FUMA. The 93 lead SNPs were selected from ∼ 800 SNPs that mapped at 89 risk loci (r^2^ ≥ 0.6 to define independent variants, default setting). This resulted in an overall Bonferroni correction of 5.62e-04 (=0.05/89 risk loci).

## Genetic correlation

LD score regression was applied to the summary statistics of the meta-analyses to calculate the genetic correlation between GSD, GSND and traits previously reported to be associated to GS, namely schizophrenia (SCZ, (Ripke et al., 2014)), bipolar disorder (BIP, (Mullins et al., 2021)), overall fracture risk (Trajanoska et al., 2018), ADHD (Demontis et al., 2019), autism spectrum disorder (ASD,(Anney et al., 2017)), coronary artery disease (CAD, (Schunkert et al., 2011)), Alzheimer’s disease (AD, (Jansen et al., 2019)), heel bone mineral density (HBD, with right and left foot) and heart attack (http://www.nealelab.is/uk-biobank). We applied Bonferroni correction to account for multiple tests, resulting in a significance threshold of 0.05 / (2 GS phenotypes x 10 associations tested) = 0.0025.

All analyses were restricted to the confidently imputed common SNPs from HapMap Phase III (1,025,494 SNPs), to avoid potential confounding effects arising from imputation quality.

## Gene-based and gene-set enrichment analysis

Gene-based association analysis and Gene-Set Enrichment Analysis (GSEA) were carried out in MAGMA (v.1.6) as implemented in FUMA (v1.3.5e). The MHC region was removed from the analysis. For the gene-based analyses, SNPs were mapped against the 18,874 protein-coding genes and associations were tested using multiple linear principal components regression for GSD and GSND. The Bonferroni-corrected threshold used to determine statistical significance for gene-based analysis was 0.05/18,874 = 2.65e-06. GSEA was conducted for all the pre-defined gene set lists from Gene Ontology (GO).

## Polygenic risk score analyses

PRS analyses were performed with PRSice 2.3.3 (Choi & O’Reilly, 2019) for GSD and GSND in the ALSPAC cohort. Summary statistics were selected from GWAS that presented a SNP-h^2^ > 0.04, including SCZ, AD, BIP, overall fracture risk, ADHD, ASD, CAD, HBD right, HBD left. From the base GWAS datasets, SNPs with MAF > 0.01 and INFO > 0.9 were filtered and then clumped (LD r^2^ ≥ 0.1 within a 250kb window). Only overlapping SNPs between the base dataset and the data in ALSPAC were retained. Age and sex were used as covariates in the analyses. A total of 9 base GWAS datasets entered the PRS analysis. We applied Bonferroni correction, resulting in a significance threshold of 0.05 / (2 target phenotypes * 9 base GWAS) = 2.77e-03. PRS analyses were run with different *p* values thresholds (0.001, 0.05, 0.1, 0.2, 0.3, 0.4, 0.5, 1) and results reported for the best *p-*value threshold (explaining the largest amount of phenotypic variation). Data preparation was performed with R v4.0.0. All analysis code is available on Github (https://github.com/fabbondanza/grip_paper).

## Results

### The grip strength phenotypes

GSD and GSND were normally distributed in both the ALSPAC and Raine Study cohorts (Figure 1). As expected from a previous study conducted in ALSPAC (Buenaventura Castillo et al., 2020), males had higher scores than females (Table 1) and scores were positively correlated with age (ALSPAC: GSD *r*_(5454)_ = 0.14, *p* < 2e-16; GSND *r*_(5454)_ = 0.13, *p* < 2e-16; Raine Study: GSD r_(df=1160)_ =0.097, *p* = 8.76E-04; GSND r_(df=1160)_=0.103, *p* = 4.41E-04; Figure S1). GSD and GSND were strongly correlated both in the ALSPAC (*r*_(5454)_ = .92, *p* < 2e-16) and in the Raine Study (r_(df=1160)_=0.87, p < 2.2e-16) cohorts.

**Figure 1:**
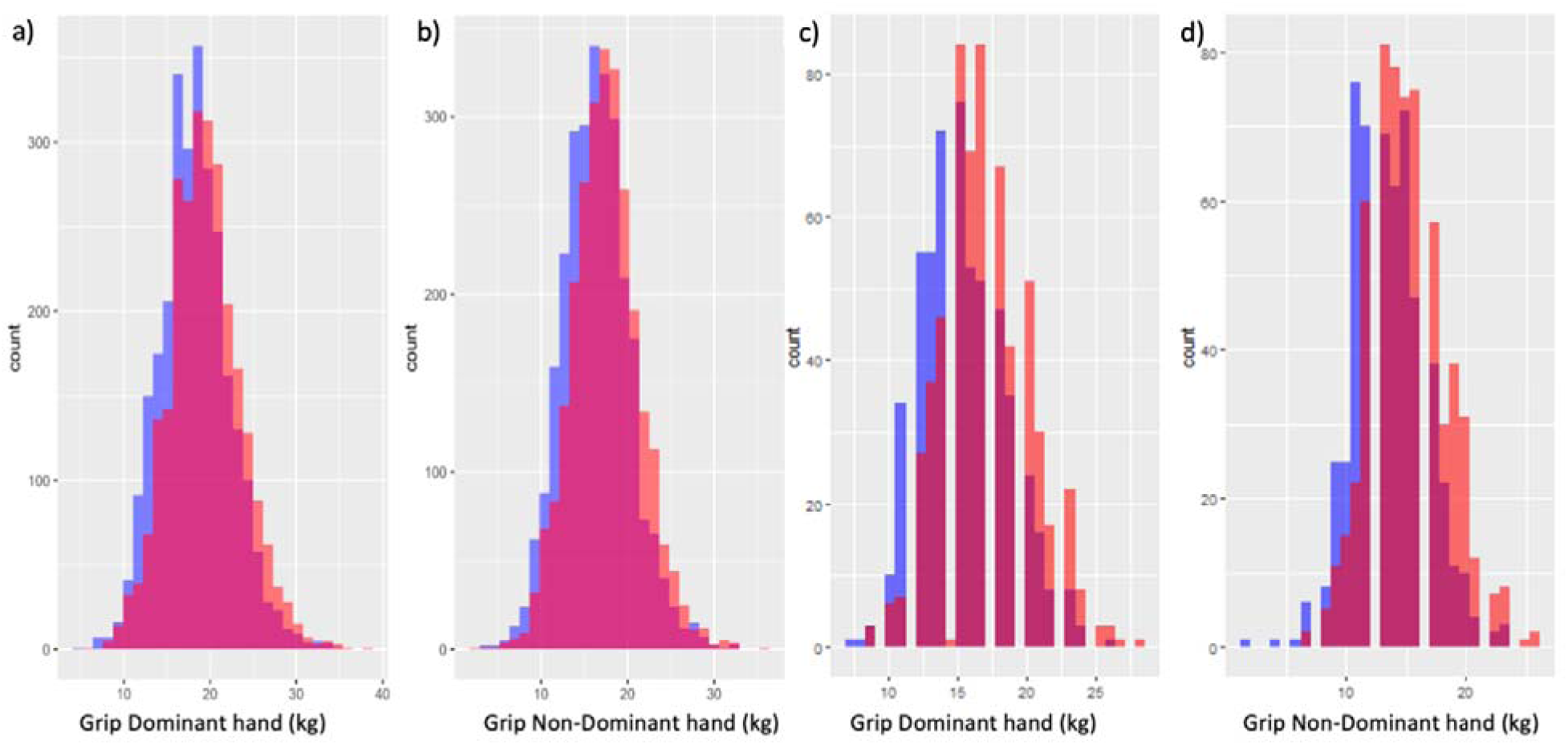
Distribution of GSD and GSND scores stratified by sex in ALSPAC (a and b) and in the Raine Study (c and d). Females in purple and males in orange.

### Heritability estimates and genetic correlation

SNP-*h*^2^, computed with BOLT-REML, was 0.393 (95% CI 0.268-0.518) for GSD and 0.41 (95% CI 0.288-0.532) for GSND in the ALSPAC sample. Estimates with LDSC, for a direct comparison with previous studies, were SNP-*h*^2^ = 0.283 (95% CI 0.095-0.471) and SNP-*h*^2^ = 0.384 (95% CI 0.184-0.584). Similar results were observed for LDSC estimates in the meta-analysis of both cohorts: SNP-*h*^2^ = 0.248 (95% CI 0.097-0.399), SNP-*h*^2^ = 0.299 (95% CI 0.142-0.456). The SNP-*h*^2^ for the non-dominant hand tended to be higher compared to the dominant with both methods, but confidence intervals overlapped.

GSD and GSND showed an almost perfect genetic correlation (r_g_ = 0.99, SE = 0.012) in ALSPAC using BOLT-REML.

### GWAS

Initially, we conducted GWAS for GSD and GSND in the ALSPAC cohort only (Supplementary Tables S1-S2, Supplementary Figure S1). No individual marker reached genome-wide statistical significance.

The strongest association observed for GSND was rs2968991 (*p* = 7.1e-08) at the long non-coding RNA [lncRNA] *LINC02465* (Supplementary Table S2). A different marker (rs2940274) at the same locus on chromosome 4 was among the top hits for GSD. Other markers among the top hits for both GSD and GSND included the *HOXB3* locus (chromosome 17) which was previously found associated with grip strength in UK Biobank (Willems et al. 2017). More than double the number of markers (434 v 197) showed *p* < 1e-04 for GSDN compared to GSD, explaining the higher SNP-h^2^ observed for GSND compared to GSD with LDSC estimates. Results for the Raine Study alone are reported in Supplementary Figure S2 and Supplementary Tables S3-S4.

The ALSPAC summary statistics were meta-analysed with the Raine Study summary statistics (Figure 2 and Table 2; Supplementary Figures S4-S5; and Supplementary Tables S5-S12). The genomic inflation factors in the meta-analyses (λ_GSD_l1=l11.040, λ_GSND_l1=l11.044) suggested significant inflation of *p*-values, but the intercept from LD-score regression (intercept_GSD_ = 1.01, intercept_GSND_ = 1.00) suggested that this was due to polygenicity rather than population stratification (see QQ plot in Supplementary Figure S3). The rs2968991 marker in *LINC02465* reached statistical significance for GSND and was among the top hits for GSD (Table 2, Supplementary Figure S4). Markers among the strongest associations for both GSD and GSND included rs2555111 in *HOXB3*, rs13069868 in *SLC9A9* and rs66461687 in *DSCAM*.

**Figure 2.**
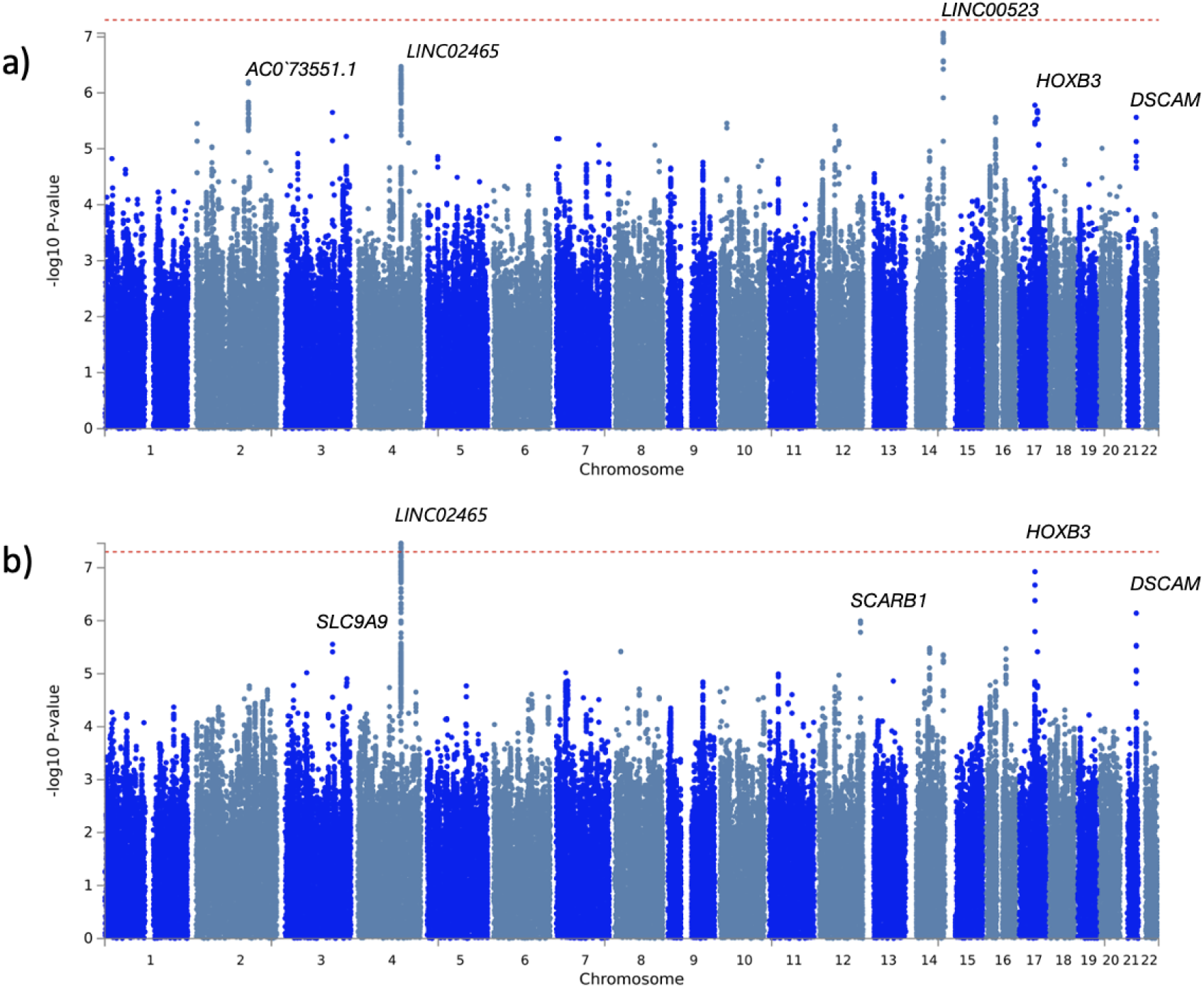
Manhattan plots for a) GSD and b) GSND in the meta-analysed sample. See Supplementary Figure S3 for QQ plots and Supplementary Figure S4 for the regional plot of the top associated locus at chromosome 4.

**Table 2.** Top associated markers in the meta-analyses.

| Trait | Location | Marker | Closest Gene | MA<br>F | Effect<br>allele | Beta | P-value | Allelic<br>direction * |
| --- | --- | --- | --- | --- | --- | --- | --- | --- |
| **GSD** | **14:101101813** | **rs78777886** | **LINC00523** | **0.06** | **T** | **-0.19** | **8.53e-08** | -- |
| **GSD** | **4:130878818** | **rs2968991** | **LINC02465** | **0.39** | **A** | **-0.08** | **3.38e-07** | -- |
| GSD† | 2:156919650 | rs111991299 | NBAS | 0.06 | T | -0.17 | 6.43e-07 | -- |
| **GSD†** | **17:46648899** | **rs2555111‡** | **HOXB3** | **0.47** | **T** | **0.08** | **1.66e-06** | ++ |
| **GSD†** | **17:54258240** | **rs6505044** | **ANKFN1** | **0.45** | **A** | **0.08** | **2.07e-06** | ++ |
| **GSD†** | **3:142972276** | **rs13069868** | **SLC9A9** | **0.26** | **T** | **-0.09** | **2.23e-06** | -- |
| **GSD** | **21:42084895** | **rs66461687** | **DSCAM** | **0.08** | **A** | **-0.15** | **2.74e-06** | -- |
| GSD† | 16:27825858 | rs1644605 | GSG1L | 0.40 | T | 0.08 | 2.75e-06 | ++ |
| GSD† | 10:21572854 | rs971015 | LUZP4P1 | 0.28 | T | 0.09 | 3.50e-06 | ++ |
| GSD† | 2:2905398 | rs34838742 | AC019118.2 | 0.19 | T | 0.10 | 3.53e-06 | ++ |
| GSD† | 12:49331240 | rs11547436 | ARF3 | 0.08 | A | 0.15 | 3.92e-06 | ++ |
| **GSND** | **4:130878818** | **rs2968991** | **LINC02465** | **0.39** | **A** | **-0.09** | **3.43e-08** | -- |
| **GSND†** | **17:46648899** | **rs2555111‡** | **HOXB3** | **0.47** | **T** | **0.09** | **1.18e-07** | ++ |
| **GSND** | **21:42084895** | **rs66461687** | **DSCAM** | **0.08** | **A** | **-0.16** | **7.17e-07** | -- |
| GSND | 12:125338529 | rs10773112‡ | SCARB1 | 0.29 | T | -0.09 | 1.01e-06 | -- |
| **GSND†** | **3:142972276** | **rs13069868** | **SLC9A9** | **0.26** | **T** | **-0.09** | **2.78e-06** | -- |
| GSND | 14:60530107 | rs61994053‡ | LRRC9 | 0.13 | T | 0.11 | 3.27e-06 | ++ |
| GSND† | 16:58684945 | rs151818‡ | SLC38A7 | 0.38 | C | -0.08 | 3.36e-06 | -- |
| GSND | 8:18993252 | rs112011071 | RP11-1080G15.1 | 0.06 | A | 0.17 | 3.79e-06 | ++ |
| **GSND†** | **17:54259113** | **rs741128** | **ANKFN1** | **0.49** | **A** | **-0.08** | **3.85e-06** | -- |
| **GSND** | **14:101101462** | **rs79742354** | **LINC00523** | **0.06** | **A** | **-0.16** | **4.41e-06** | -- |

Loci reported in the top variants for both GSD and GSND are in bold
\*Refers to the direction of the association for ALSPAC and Raine Study, respectively
†Refers to loci found to be associated with GS traits in previous literature (see Replication analysis and Supplementary Table S14)
‡Refers to variants that showed eQTL effects in GTEx v8 (See Supplementary Tables S7, S11)

The rs2555111 variant in *HOXB3* showed a strong eQTL effect (lowest p-value across multiple tissues: *p* = 1.40e-62), with each copy of the allele associated with higher grip strength (T allele), upregulating *HOXB-AS1, HOXB2*, *HOXB3*, *HOXB6* and *HOXB7* across multiple tissues (for the full eQTL results, see supplementary tables S7, S11).

The top associated variants were annotated for previously reported GWAS associations which included body mass index (*LINC02465*) and heel bone mineral density (*ANKFN1*) (Supplementary Tables S8, S12). We then conducted a replication analysis for 93 lead SNPs (Supplementary Table S13) previously reported to be associated with grip strength. In total, 22 SNPs showed a statistically significant association corrected for multiple testing (*p* < 5.62e-04) (Table 2; Supplementary Table S14). Two markers, rs2555111 in *HOXB3* and rs13069868 in *SLC9A9,* showed association with both GSD and GSND.

No significant results were found for gene-based analyses and gene set enrichment analyses (GSEA) for GSD and GSND in the meta-analysed data (Supplementary Figure S5, supplementary Table S6, S10).

### Genetic correlation and Polygenic risk score analyses

Genetic correlations were tested for psychiatric (ADHD, BIP, ASD, SCZ, AD) and non-psychiatric (heel bone density with the left [HBD left] and right foot [HBD right], CAD, heart attack, overall fracture risk) traits (Figure 3, Supplementary Table S15). We found a statistically significant positive correlation with CAD for both GSD (r_g_ = 0.20, *p* = 8e-04) and GSND (r_g_ = 0.19, *p* = 1e-04) that were consistent with previous reports for GS in an adult cohort (r_g_ = 0.19) (Jones et al. 2021). Nominally significant correlations were observed for self-reported history of heart attack (r_GSND_ = 0.081 [SE = 0.037], *p*_GSND_ = 0.03) and negative correlations with ADHD (r_GSD_ = –0.064 [SE = 0.030], *p*_GSD_ = 0.036), ASD (r_GSD_ r = –0.071 [SE = 0.033], *p*_GSD_ = 0.033) and overall fracture risk (r_GSD_ = –0.129 [SE = 0.045], *p*_GSD_ = 0.0037; r_GSND_ = –0.097 [SE = 0.039], *p*_GSND_ = 0.012).

**Figure 3:**
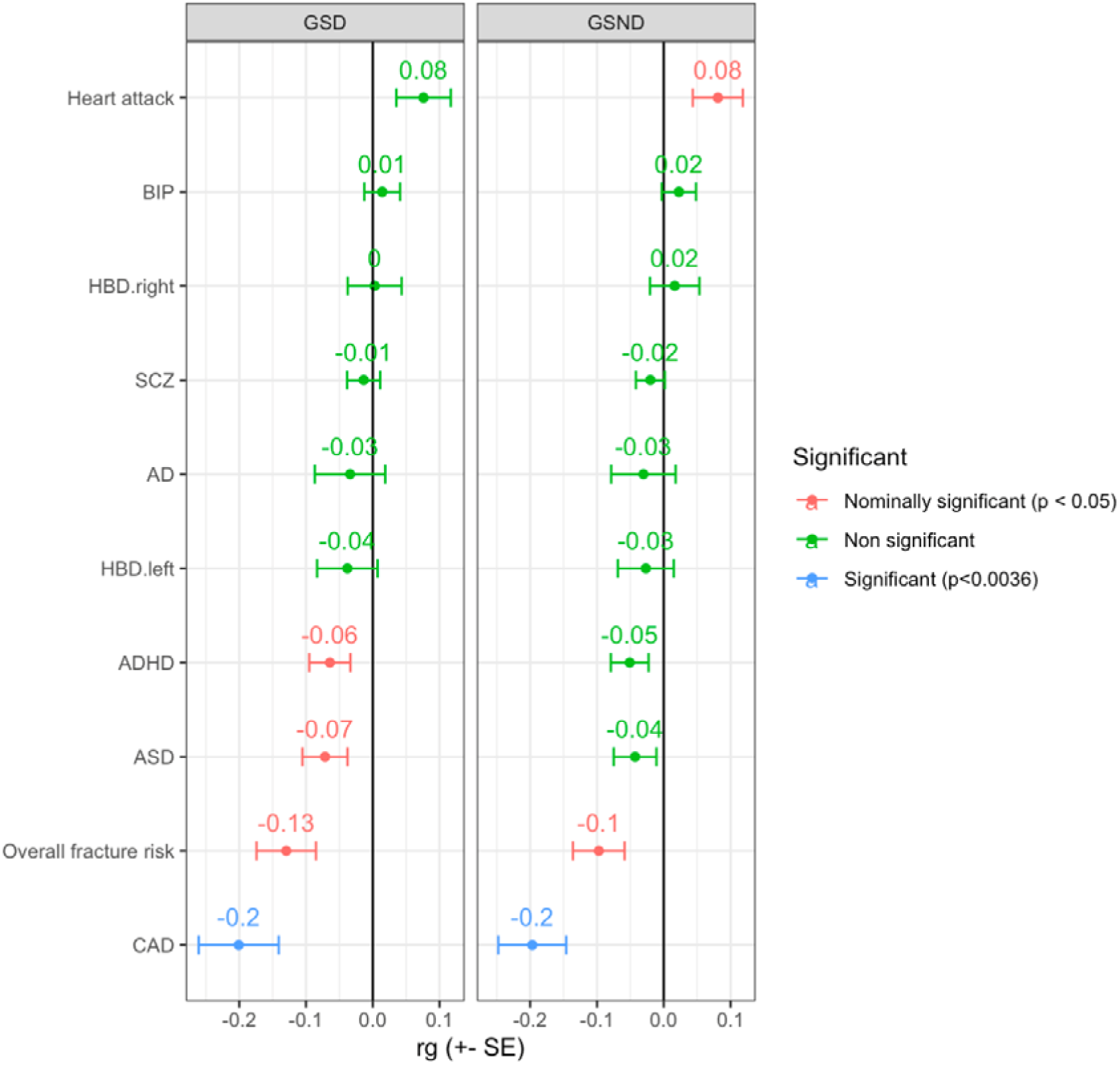
G**e**netic **correlation (rg) analysis for GSD and GSND**. SCZ = Schizophrenia, BIP = Bipolar disorder, ASD = Autism spectrum disorder, HBD.right = Heel bone density with the right foot, HBD.left = Heel bone density with the left foot, AD = Alzheimer’s disease.

We then conducted PRS analysis for the same set of traits after excluding heart attack due to the low SNP-h^2^ (SNP-h^2^ < 0.04) (Table 3, Supplementary Table S16). Overall fracture risk was negatively associated with both GSD and GSND (p < 2.77e-03). The standardised beta coefficients indicate that an increase of 1 SD in PGS is associated with a decrease of 0.04 SD in both GSD and GSND.

**Table 3:** Summary of the PRS analyses.

| Target trait | Base GWAS | PRS R<sup>2</sup> (%) | Full R<sup>2</sup> (%) | Beta (SE) | p-value |
| --- | --- | --- | --- | --- | --- |
| **GSD** | **Overall fracture risk** | **0.16** | **3.75** | **-0.04 (0.01)** | **2.76E-03** |
| GSD | ADHD | 0.14 | 3.72 | 0.04 (0.01) | 5.85E-03 |
| GSD | HBD left | 0.10 | 3.69 | 0.03 (0.01) | 1.70E-02 |
| GSD | HBD right | 0.06 | 3.65 | 0.02 (0.01) | 7.14E-02 |
| GSND | ADHD | 0.15 | 3.68 | 0.04 (0.01) | 3.87E-03 |
| **GSND** | **Overall fracture risk** | **0.18** | **3.71** | **-0.04 (0.01)** | **1.69E-03** |
| GSND | HBD left | 0.11 | 3.64 | 0.04 (0.01) | 1.31E-02 |

Results are shown for the best $p$ value thresholds (highest R<sup>2</sup>). Only nominally significant results are reported ( $p < 0.05$ ). Betas are standardised. Statistically significant associations are in bold ( $p < 2.77\text{e-}03$ ). Full results are shown in Supplementary Table S16.

## Discussion

We report the first GWAS for grip strength measures in children. Previous genetic studies for GS were conducted predominantly in adult cohorts and focussed on measures of maximal strength, typically for the dominant hand. GS measures assessed in adults are likely to be confounded by environmental factors, linked to lifestyle, type of work or recreational activities with a stronger effect on the dominant hand because of more training.

Consistent with our reasoning, GSND showed a trend a higher SNP-*h*^2^ compared to GSD when estimated with BOLT-REML (0.39 v 0.41) and especially with LDSC (0.28 v 0.38; ALSPAC sample only). The difference in the latter case could have resulted from a lower number of SNPs for GSD compared to GSND passed the p-value cut-off to be included in the LDSC calculation. The SNP–*h2* observed in our study were higher compared to estimates observed in adults which ranged from 13% (Tikkanen et al., 2018) to 24% (Willems et al., 2017) and were as low as 4% in older (>65 years old) individuals (Jones et al., 2021). Our results in combination with observations from the literature suggest that the heritability of grip strength decreases in older age because of environmental factors (Mitchell et al., 2012).

Despite the smaller sample size of our study compared to analyses in adult cohorts, we both replicated previously reported loci and reported a novel statistically significant association, demonstrating the advantages of analysing cohorts of children. We anticipate that analyses in larger cohorts of children will lead to additional genetic associations. The novel association was detected for GSND at the *LINC02465* locus on chromosome 4. The *LINC02465* long non-coding RNA has previously been found to be associated with body mass index and body size (Davies et al., 2018; Pulit et al., 2019; Sakaue et al., 2021). The same locus ranked among the top associations also for GSD.

Different genes, including *LINC00523* (top association for GSD), *HOXB3, SLC9A9, ANKFN1* and *DSCAM* were among the top associations for both GSD and GSND. Given the strong correlation between GSD and GSND, such consistency it is not surprising, nevertheless it represents an internal control for our analyses. The reliability of our findings is further supported by the replication of 22 out of 93 associations previously reported in the literature, including for the *HOXB3, SLC9A9, and ANKFN1* genes observed in top associations for both GSD and GSND.

The *HOXB3* association is of particular interest because it was underpinned by the same SNP (rs2555111) which also shows strong eQTL effects regulating multiple HOXB genes (e.g., *HOXB-AS1, HOXB2, HOXB3, HOXB6, HOXB7*) across multiple tissues, including muscle-skeletal tissue.

When testing for genetic correlations risk for CAD showed a statistically significant signal consistent with what reported by Jones and colleagues 2021 in adults (Jones et al., 2021).

PRS analyses showed an association between risk for overall fracture and low GSD and GSND. Although not statistically significant, we observed a pattern of association between stronger heel bone density and higher GS scores. The PRS results for overall fracture risk and heel bone density are in line with associations observed at phenotypic level (Cheung et al., 2012; Kärkkäinen et al., 2008).

In conclusion, by performing the first GWAS for grip strength in children, we both report novel findings as well as replicating associations previously reported in cohorts of adults but much larger in sample size. We hypothesise that analyses in children are less confounded and the availability of larger children cohorts has the potential to advance our understanding of GS genetics which, in turn, might predict a number of health outcomes.

## Data Availability

The ALSPAC study website contains details of all the data that is available through a fully searchable data dictionary (http://www.bris.ac.uk/alspac/researchers/data-access/data-dictionary/). The data access policy is described here https://www.bristol.ac.uk/alspac/researchers/access/.
The Raine Study website contains details of all the data that is available https://rainestudy.org.au/information-for-researchers/available-data/. The data access policy is described here https://rainestudy.org.au/information-for-researchers/project-application-process/

https://github.com/fabbondanza/grip_paper

## Acknowledgment

The authors are grateful to all the families who took part in this study, the midwives for their help in recruiting them, and the whole ALSPAC team, which includes interviewers, computer and laboratory technicians, clerical workers, research scientists, volunteers, managers, receptionists and nurses.

The authors are grateful to the Raine Study participants and their families, and to the Raine Study team for cohort coordination and data collection. The authors gratefully acknowledge the NHMRC for their long-term funding to the study over the last 30 years and also the following institutes for providing funding for Core Management of the Raine Study: The University of Western Australia (UWA), Curtin University, Women and Infants Research Foundation, Telethon Kids Institute, Edith Cowan University, Murdoch University, The University of Notre Dame Australia and The Raine Medical Research Foundation. This work was supported by resources provided by the Pawsey Supercomputing Centre with funding from the Australian Government and Government of Western Australia.

Finally, the authors thank all the studies and databases that made GWAS summary data available.

## URL

PGC summary statistics – https://www.med.unc.edu/pgc/results-and-downloads/

Neale’s Lab UK Biobank summary statistics – http://www.nealelab.is/uk-biobank

Summary statistics for CAD – http://www.cardiogramplusc4d.org/data-downloads/

