## Supplementary Figures for "A GWAS for grip strength in cohorts of children – advantages of analysing young participants for this trait"

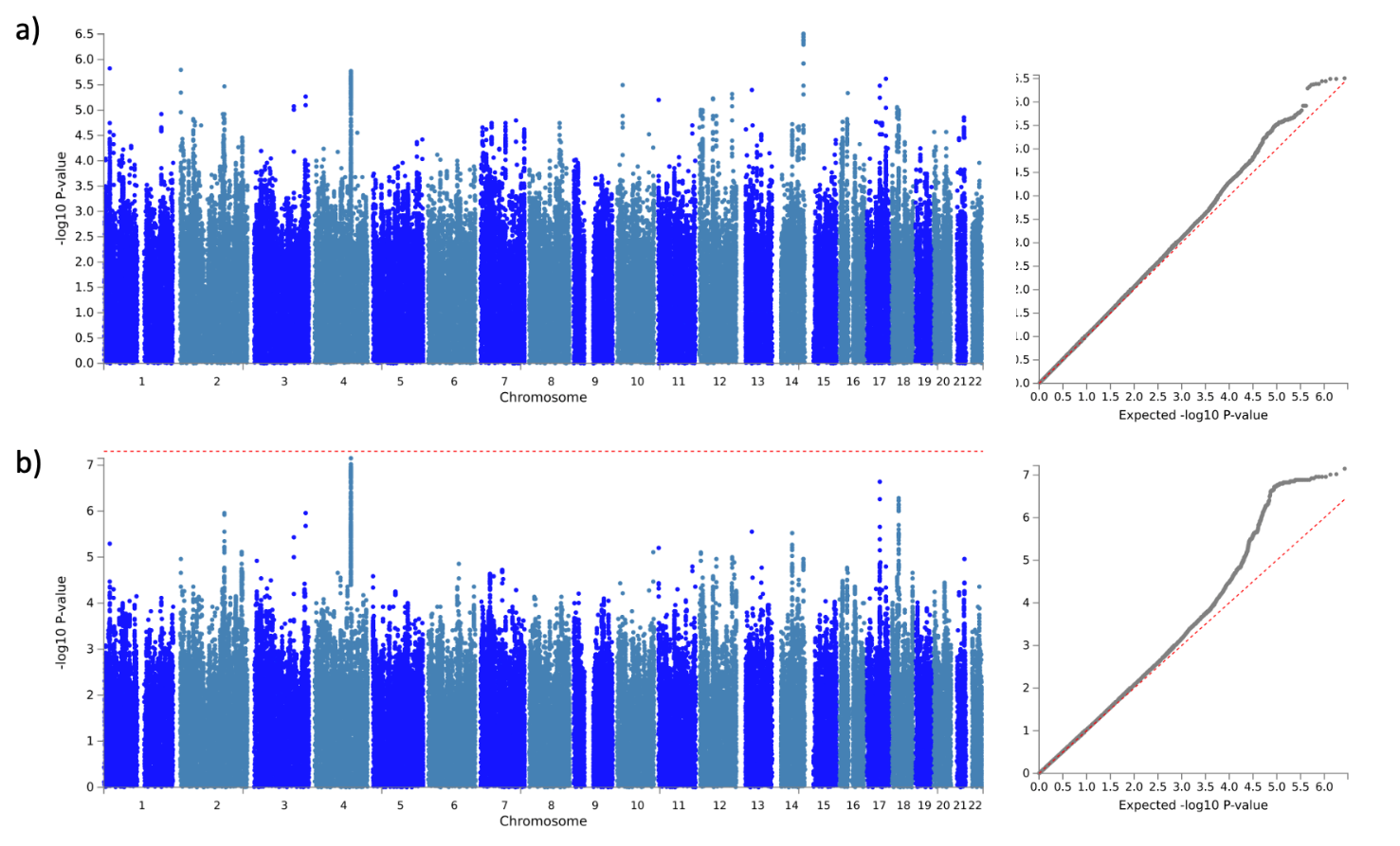
**Supplementary Figure S1. Manhattan and** QQ-plots for a) GSD and b) GSND in the ALSPAC cohort.

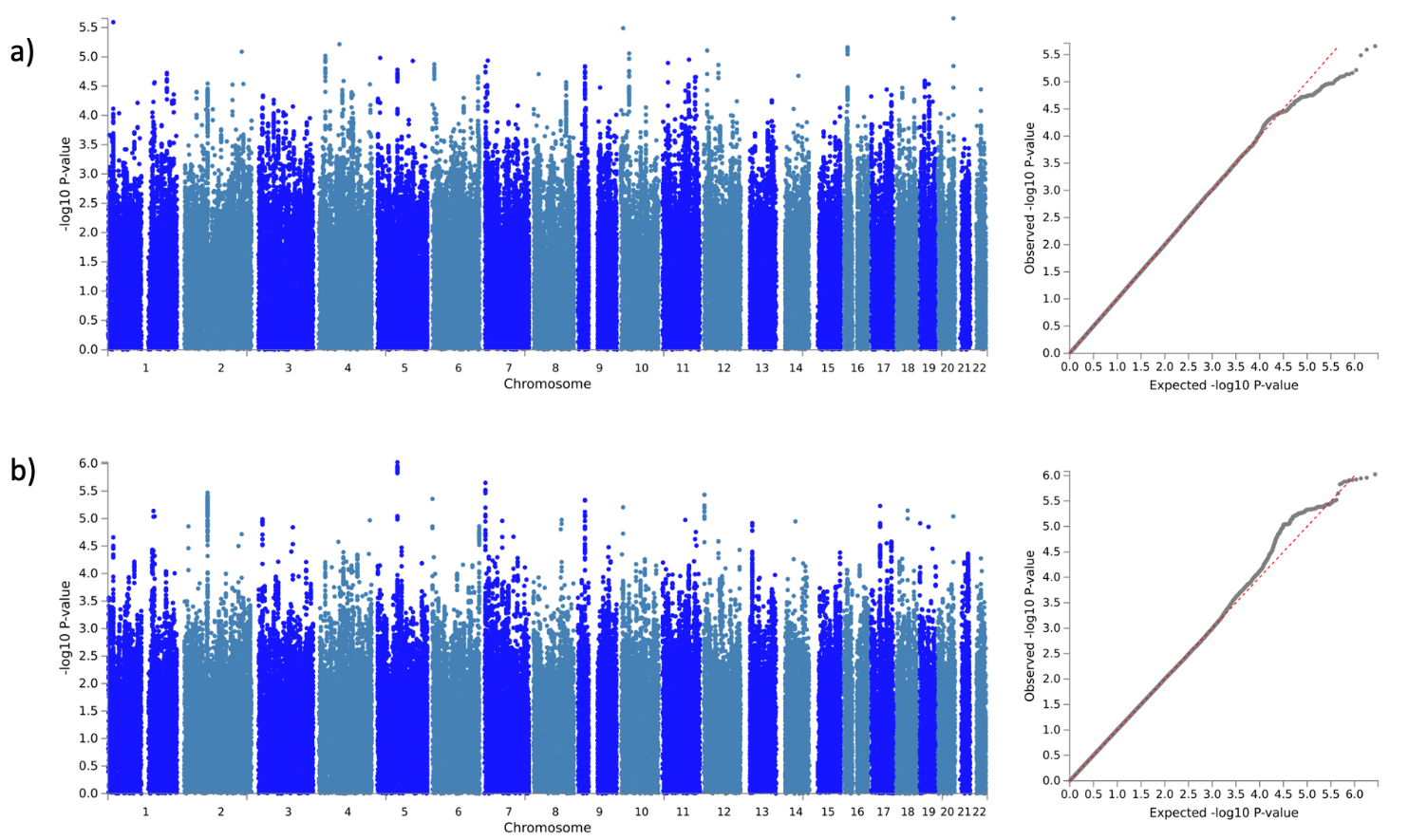

**Supplementary Figure S2:** GWAS and QQ-plot for a) GSD and b) GSND in the Raine Study

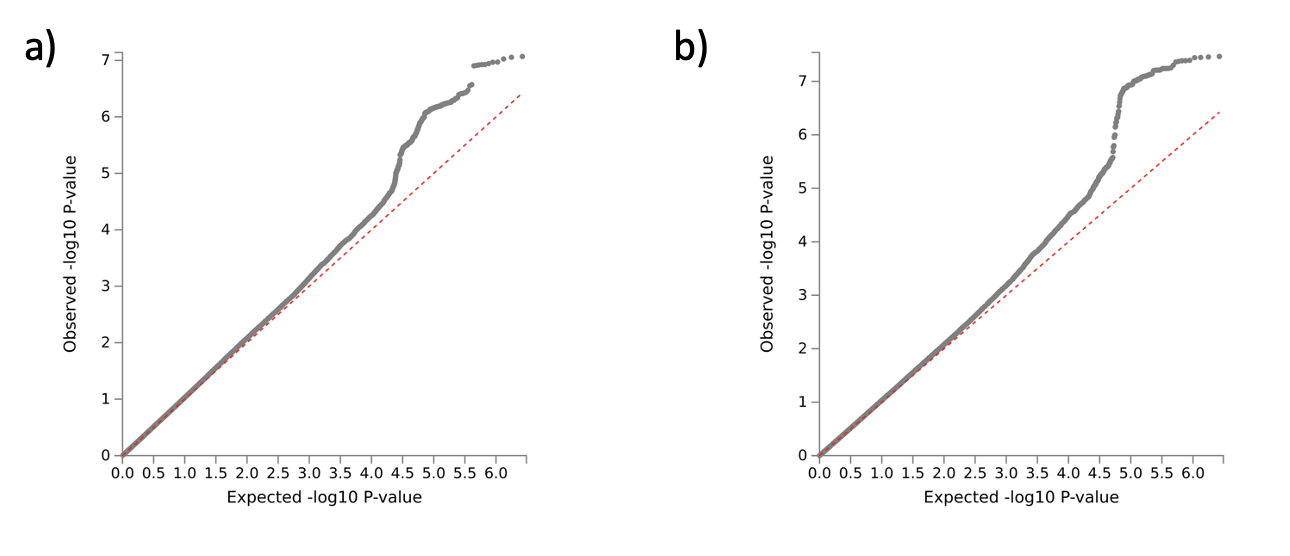

**Supplementary Figure S3:** QQ-plot for a) GSD and b) GSND in the meta-analysed sample.

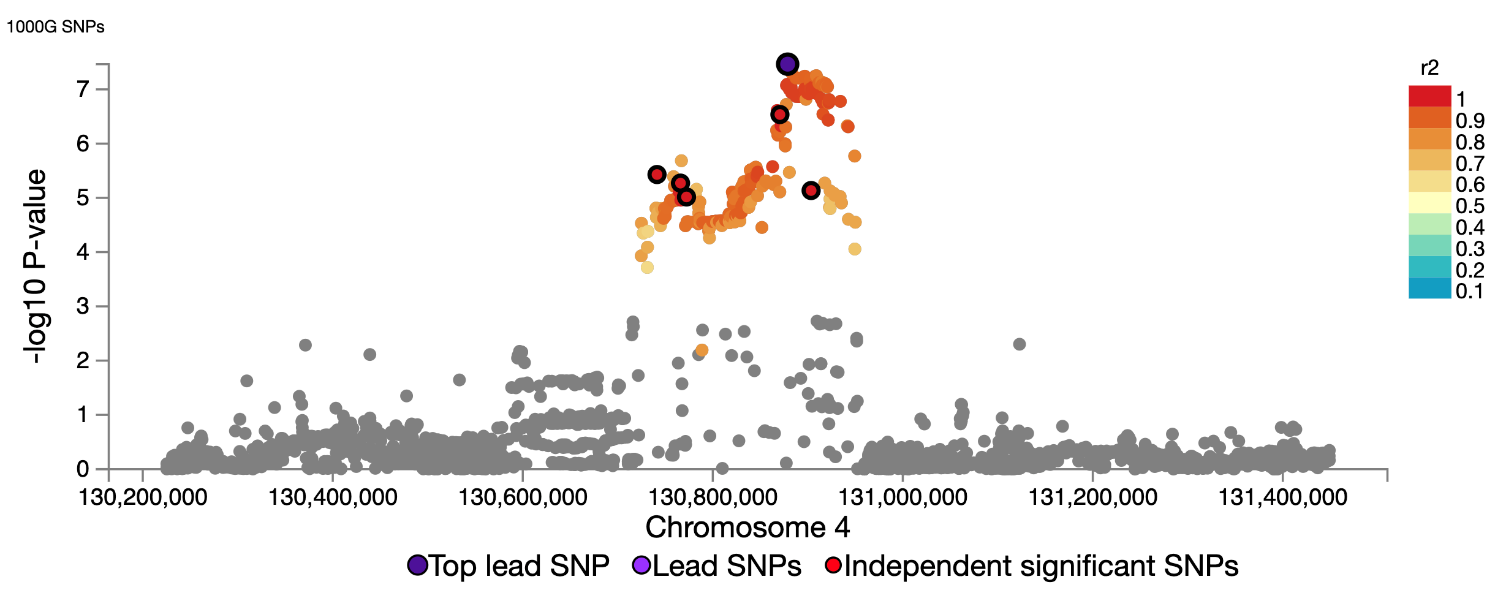

**Supplementary Figure S4:** Regional plot for rs2968991 in the GSND meta-analysis.

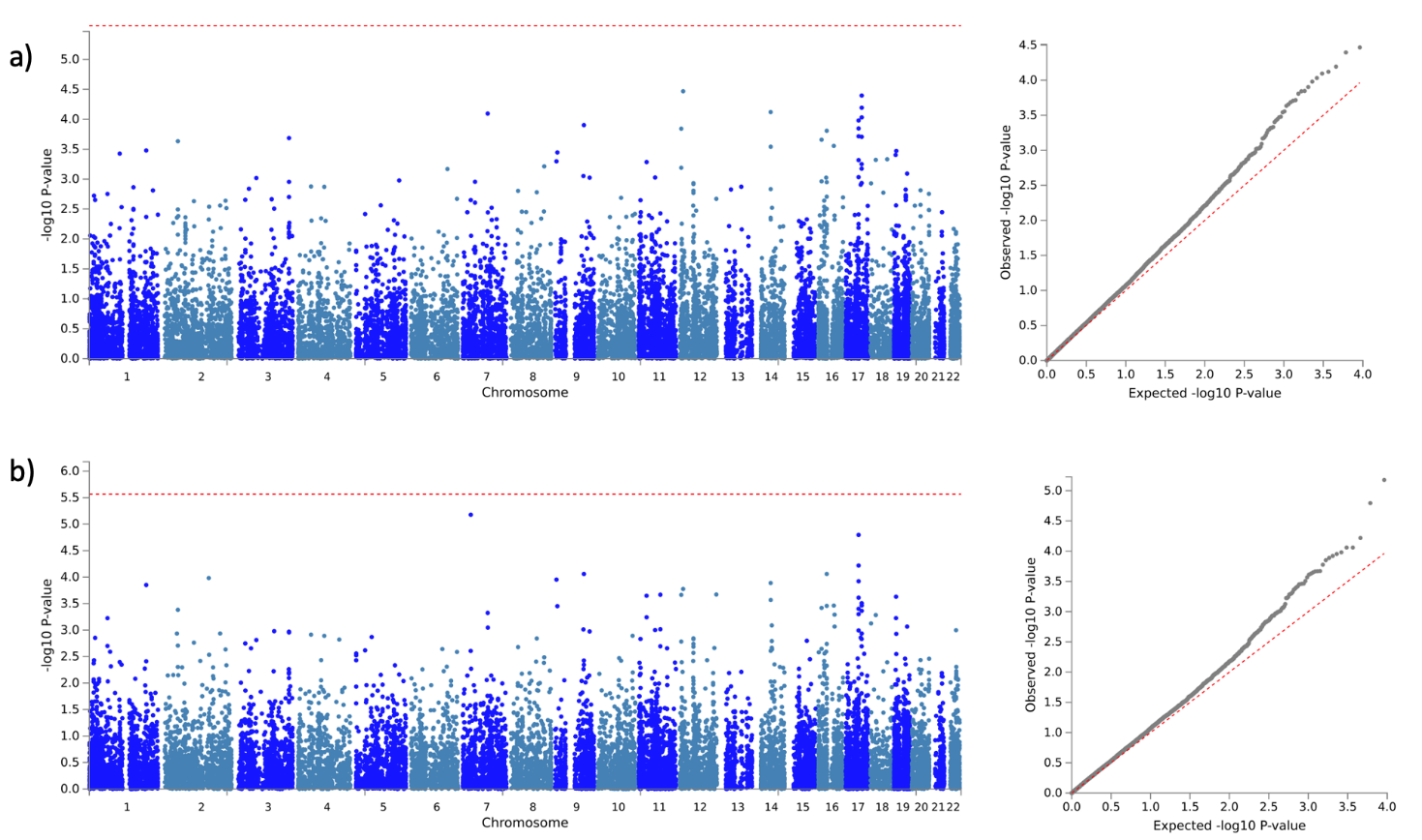
**Figure S5**: Gene-based meta-analysis Manhattan plots for a) GSD and b) GSND in the meta-analysed sample
